# Effect of a mobile health intervention optimised with artificial intelligence on blood pressure and other cardiovascular risk factors in adults with high blood pressure: Rationale and design of My Intelligent Cardiac Assistant (MICArdiac) Randomised Controlled Trial

**DOI:** 10.1101/2025.10.14.25337964

**Authors:** Liliana Laranjo, Edel O’Hagan, Harry Klimis, Simone Marschner, Clara K Chow

## Abstract

**Introduction:** Control of blood pressure (BP) continues to be a challenge globally. Clinical trials have shown home BP monitoring and text-message interventions to lower BP. Integrating these to personalise supportive messaging in response to changing BP and activity through leveraging artificial intelligence (AI) could improve BP control. The aim of this trial is to examine the impact on BP, compared to a text-message only program, of the My Intelligent Cardiac Assistant (MICArdiac) program. MICArdiac is a 6-month program comprising tracking of BP, physical activity and heart rate informing a content stream of AI-driven personalised messages to encourage BP self-management, as-necessary medical review to up-titrate medicines and cardiovascular preventative behavioural change.

**Methods and analysis:** MICArdiac is a prospective randomized open-blinded endpoint randomised controlled trial with 1:1 (intervention:control) allocation and active control. Individuals aged 35 years old or above with high BP are randomised to the MICArdiac program or to a text-message cardiovascular education program. Recruitment started in primary care and hospital clinics in Western Sydney and moved to decentralised direct-to-community recruitment in the Australian Capital Territory, New South Wales, and Victoria. Randomisation and allocation concealment occur via a secure web-based system. The primary outcome is mean daytime systolic BP measured by 24-hour Ambulatory Blood Pressure Monitoring at 6 months. Primary analysis will follow intention-to-treat principles; data analysts will be blinded. Process evaluation will be conducted. The study has recruited 451 participants, giving it 94% power to detect a difference of 4 mmHg in the primary outcome.

**Ethics:** Ethics approval was obtained from Western Sydney Local Health District Human Ethics Research Committee (2021/ETH11379).

**Clinical trials registration number:** Australian New Zealand Clinical Trials Registry (Registration number: ACTRN12622000091707).

## INTRODUCTION

Cardiovascular disease (CVD) remains the largest cause of premature death and disability worldwide^1^. Much of the burden of CVD is attributable to poor BP control as well as other uncontrolled CVD risk factors.^2^ Optimising health behaviours, including lifestyle and medication adherence, is a key strategy to improve BP control and reduce CVD risk^3–9^. A range of CVD prevention interventions that provide education, health behaviour change support and thus encourage self-management of high BP have been shown to improve BP and CVD risk factors known to be associated with reduction in future cardiovascular events^4, 10^. Yet common barriers to scale-up involve access and availability, heavy resource utilisation including clinician time, and patient engagement^6, 10^.

Mobile health technologies such as text-messaging, mobile apps, and wireless monitoring devices may help overcome these barriers. A 2023 systematic review and meta-analysis found that digital interventions (smartphone apps, websites or SMS) were associated with a significant reduction in systolic (mean difference -3.62 mmHg (95% CI-5.22 to -2.02)) and diastolic (mean difference -2.45 mmHg (95% CI-3.83 to -1.07)) BP compared to control group.^11^ Heterogeneity was high and there was wide variability between the different types of app interventions: some focused only on medication adherence^12^ or on a specific lifestyle behaviour (e.g., diet)^13^. Meta-regression was not conducted to identify specific app components associated with higher effectiveness.

More recent trials evaluating standalone app interventions (i.e., without clinician involvement) for patients with high BP have found mixed results^14, 15^. A trial comparing self-monitoring of BP with app-based self-monitoring (i.e., via connected wireless BP monitor) did not find a significant difference in BP^14^, but the intervention did not provide support for lifestyle behaviour change or medication adherence. Another trial evaluated app-based self-monitoring and lifestyle behaviour change support for patients not medicated with anti-hypertensives, showing a small significant decrease (∼2 mmHg) in systolic BP compared to usual care^15^. Finally, a trial evaluating an app-based self-monitoring and lifestyle behaviour change support via conversational artificial intelligence (AI) in comparison to just app-based self-monitoring did not find a significant difference in BP^16^. However, none of the interventions in these trials provided education and support on medication adherence, nor did they support patients in seeking medication up-titration for BP control where needed.

To date, no trial has evaluated a comprehensive mobile health program to support patients with high BP self-manage. Our proposed intervention incorporates app-based monitoring of BP, heart rate and physical activity (via connected wireless devices), AI-enhanced personalised education and behaviour change support (for lifestyle behaviours and medication adherence), and prompts with letters for patients to show their GP or Cardiologist for a medication review as triggered through algorithms for non-controlled BP. Novel AI methods, including machine learning and natural language processing, have been suggested as promising approaches for more engaging and personalised self-management interventions^17–19^, but trials assessing their effectiveness in BP management are sparse^16^.

The primary objective of this trial is to assess the impact of an AI enhanced mobile health program - the <u>My</u> Intelligent <u>C</u>ardiac <u>A</u>ssistant (MICArdiac)-, compared to standard text messaging education for cardiovascular disease prevention, on daytime average ambulatory systolic BP, in participants with high BP. Secondary objectives are to determine the impact of the intervention on other BP measures and CVD risk factors, quality of life, cardiovascular knowledge, and unplanned hospitalisations, as well as conduct a process evaluation, assessing engagement, acceptability, and implementation of the intervention.

## METHODS AND ANALYSIS

### Study Design

MICArdiac is a single-blind 1:1 (intervention:control) randomised controlled trial (RCT) with 6 months of follow-up (Figure 1). The trial has been registered in the Australian and New Zealand Clinical Trials Registry (Registration number: ACTRN12622000091707) and the study protocol is reported following the SPIRIT-AI checklist^20^ for RCT protocols (Supplement 1) and the mobile health (mHealth) evidence reporting and assessment (mERA) checklist^21^ (Supplement 2).

**Figure 1:**
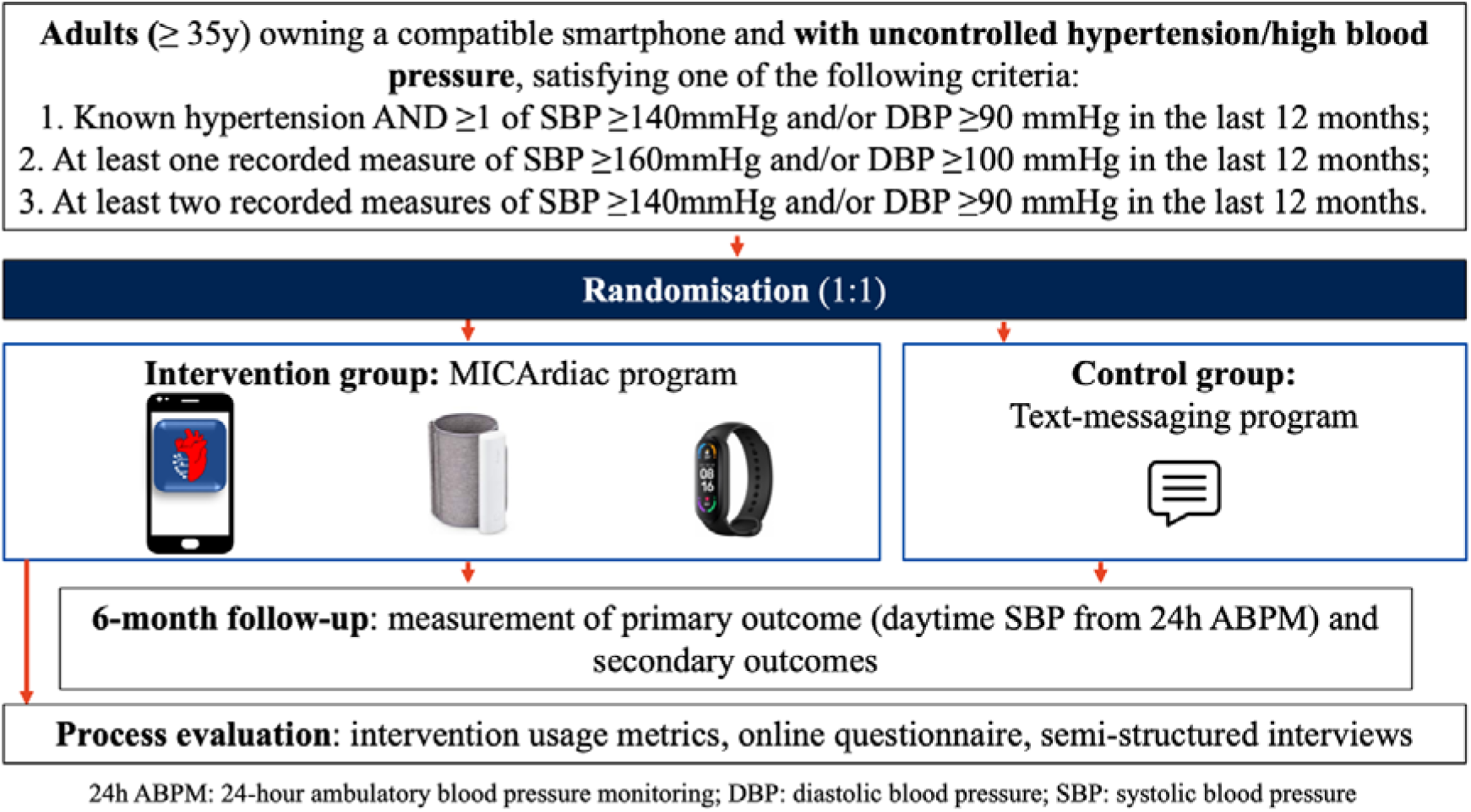
Study flow diagram

### Study setting

This study commenced recruitment in Western Sydney, NSW, Australia, with the initial sites as Westmead Hospital outpatient services and General Practice clinics in Western Sydney Primary Health Network. Recruitment was then decentralised and extended to community recruitment in New South Wales, Victoria and the Australian Capital Territory. In Australia, nine out of ten adults own a smartphone^22^.

### Participants

Potential participants are eligible to participate if they are 35 years old or above, own a compatible smartphone – iPhone 5 onwards (iOS 10 and higher) or Android (6.0 and higher), and have uncontrolled hypertension or high BP, satisfying one of the following criteria:

- Known hypertension AND at least one recorded measure of systolic BP ≥140mmHg and/or diastolic BP ≥90 mmHg in the last 12 months; OR
- At least one recorded measure of systolic BP ≥160mmHg and/or diastolic BP ≥100 mmHg in the last 12 months; OR
- At least two recorded measures of systolic BP ≥140mmHg and/or diastolic BP ≥90 mmHg in the last 12 months.

Participants will be excluded from the study if they required acute care in the preceding 30 days in relation to cardiovascular disease (e.g., heart failure, coronary heart disease, stroke, transient ischemic attack). Additional exclusion criteria include being unable to use the monitoring devices provided; being unable to complete the study procedures or follow-up; being unable to understand sufficient written English to provide informed consent; or having a concomitant illness, physical impairment, or mental condition which in the opinion of the study team/primary physician could interfere with the conduct of the study. Participant flow through the study is presented in Figure 1.

### Intervention: The MICArdiac Program

All participants randomised into the intervention arm will receive the MICArdiac program for 6 months. The MICArdiac program (Supplement 3) aims to improve BP and other cardiovascular risk factors and consists of cardiovascular education and behaviour change support through a mobile health program—MICArdiac smartphone App (Figures 2 and 3)—that is connected with wireless monitoring devices (physical activity tracker and BP monitor). After randomisation, devices are provided to intervention participants (in person or mailed) with instructions to download the MICArdiac app and setup the devices; a research assistant is available to support and troubleshoot the setup process and any emerging device issues during the study.

The MICArdiac program comprises tracking of BP, physical activity and heart rate, and a content stream of AI-driven personalised messages (delivered via mobile app or text-messages) to encourage BP self-management and as-necessary medical review to up-titrate medicines in addition to CVD prevention program content. By integrating data from wireless devices and goal-related messages to deliver personalised self-monitoring and lifestyle modification support, the program incorporates several behaviour change techniques, based on the Capability Opportunity Motivation Behaviour (COM-B) model^23, 24^ (supplement 4).

**Figure 2:**
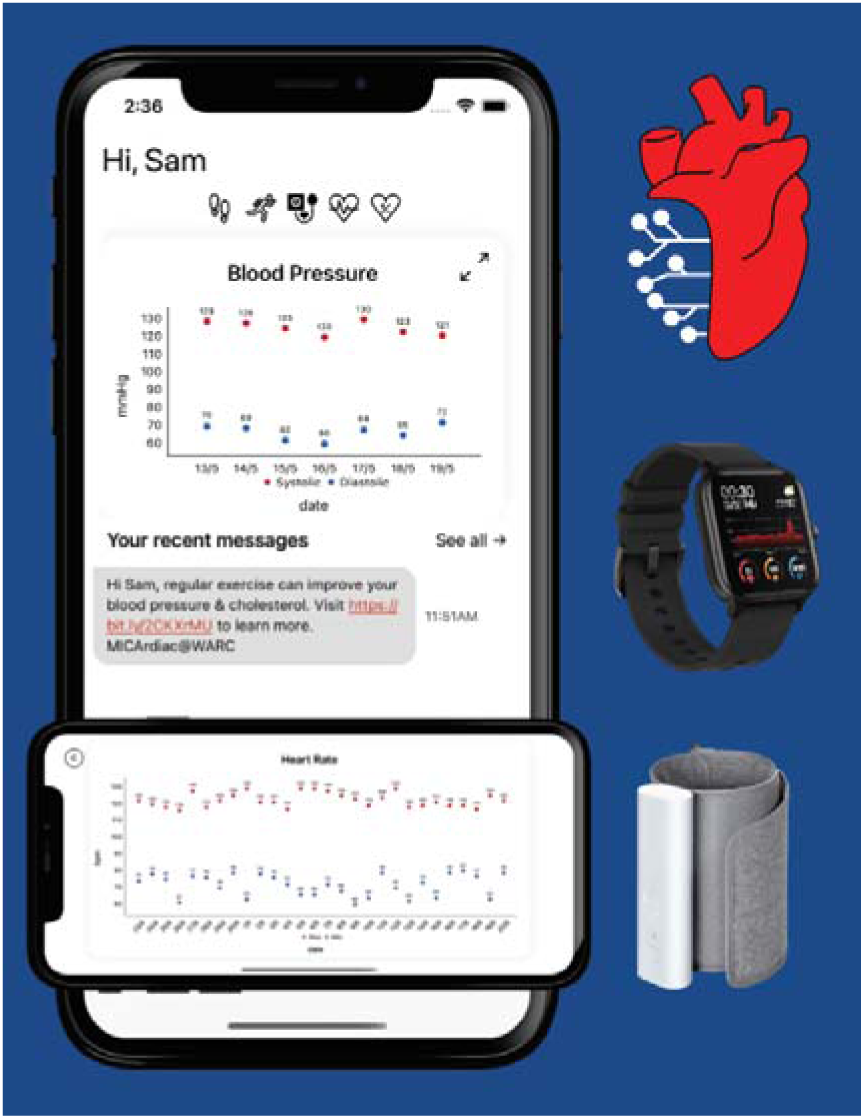
MICArdiac app and devices

**Figure 3:**
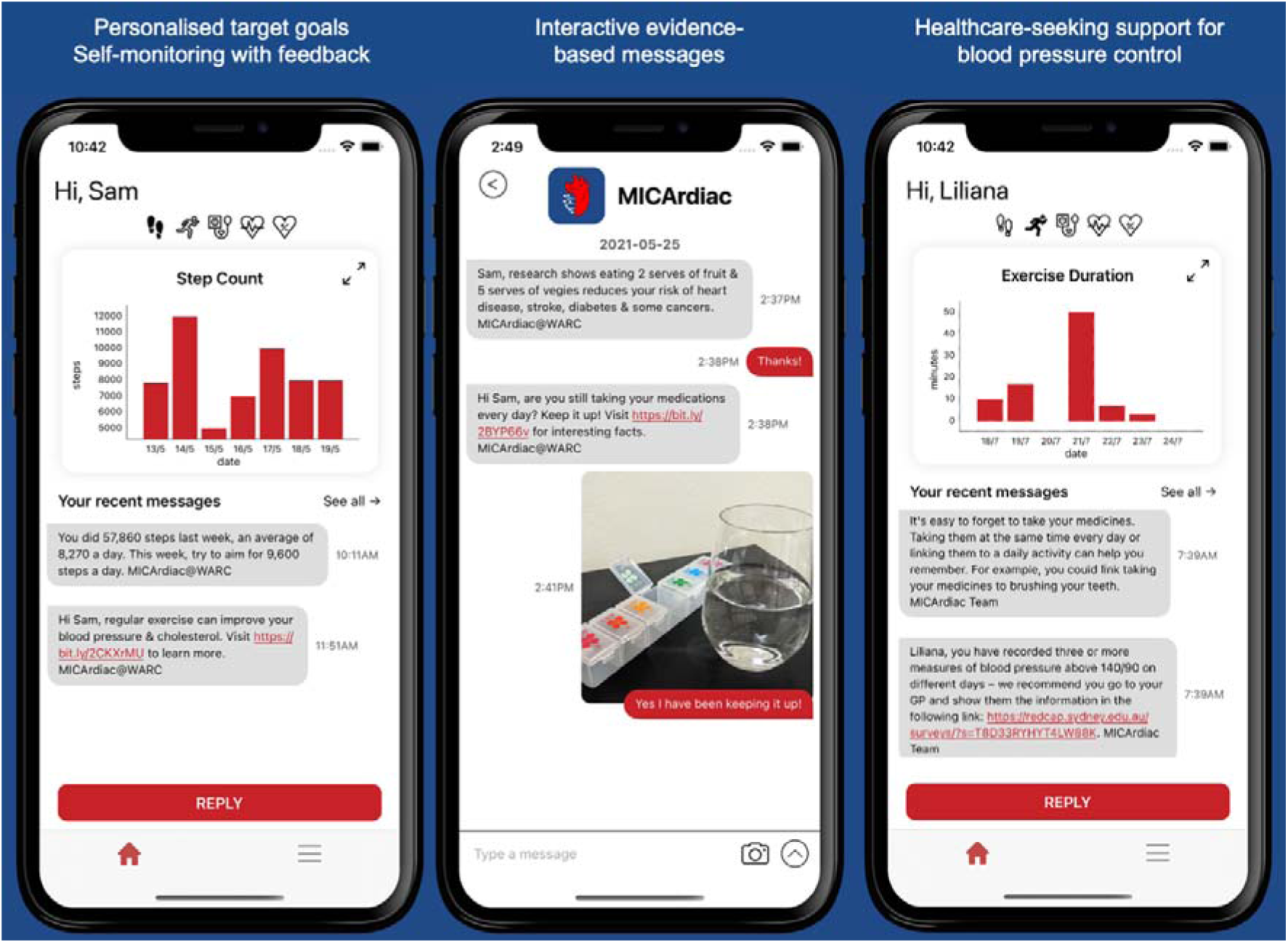
MICArdiac features

### Wireless monitoring devices

Devices include a wireless BP monitor (Withings BPM Connect, validated^25^ and recommended by the STRIDE BP, the international scientific organisation dedicated to ensuring the accuracy of blood pressure measurement^26^) and a physical activity tracker with heart rate monitoring ability (Xiaomi Mi Smart Band or a ‘Bring Your Own device’ option, depending on participant preference). Participants are encouraged to always wear the activity tracker, removing the bracelet only when required to charge it. Participants are instructed to measure their BP using the wireless device two times a week at different times of the day.

### MICArdiac app

The MICArdiac app graphically displays data collected from the wireless monitoring devices (step counts, exercise duration, BP, heart rate) and delivers personalised messages utilising data from the wireless monitoring devices, AI or threshold-based algorithms (see *messages*, below). The MICArdiac app is downloadable via the Apple and Google app stores, and it securely collects the participant’s BP measures, heart rate and physical activity measures from the smartphone’s GoogleFit app or Apple Healthkit app.

### Messages

The frequency of messages will vary between 4 and 7 messages per week. Messages can be delivered as in-app or text message (i.e. Short Message Service, SMS), depending on patient preference. The MICArdiac intervention is designed to be interactive and supportive with participants encouraged to engage with the health counsellor if they have any questions or concerns. Messages will be of three main types:

I. **Educational messages** to encourage behaviour change and self-management, drawn from a message bank of at least 5 topic streams: BP (including medication adherence), cholesterol, diet, physical activity, support and smoking (if applicable). Messages are customised using one baseline variable (smoking status) and their content and sequence are optimised by a machine learning algorithm focusing on message intent, lexical similarity, and message readability (based on participant’s health literacy, measured at baseline). The algorithm was developed based on natural language processing analysis of messages and responses from our previous RCTs (further information on the AI-enhanced message delivery protocol available in Supplement 3)^27, 28^. The content was adapted from previous trials and developed by a team of clinicians (GP, cardiologists, physiotherapists, dieticians) based on national guidelines and information from reputable sources (e.g., National Heart Foundation) and refined with consumer input (Supplement 5).
II. **Adaptive algorithm** messages containing **weekly adaptive step goals** based on data from the physical activity tracker. The adaptive algorithm first monitors the participant’s step count for one week at baseline and then it generates a goal for next week based on this number. The algorithm then generates a new goal each week based on the participant’s step count and their ability to achieve the previous week’s goal. The aim is to gradually increase participants’ step goals in achievable increments.
III. **Threshold-based messages based on data from the wireless monitoring devices.** These messages will be rule-based and one of three types: 1) messages indicating that the values of health parameters (BP, heart rate) are within the normal/recommended range and encouraging maintenance of health behaviours and management; 2) messages indicating that the values are out of the normal/recommended range and encouraging improvement of health behaviours. When BP thresholds are met these will trigger messages to prompt patients to go to their usual doctors for medical review and the message prompting this will embed a letter of referral to their doctor containing high BP information and why they need medical review. Extreme values of BP and heart rate will trigger a phone call from a health counsellor; and 3) messages reminding participants to use wireless monitoring devices, triggered by a lack of data received in the previous week.

### The MICArdiac program development

Before commencing recruitment, the MICArdiac program was evaluated in a single-arm pilot study to assess user experience and refine the intervention. Nine participants were purposively recruited to ensure diversity in age, sex, cultural background, lifestyle, and baseline cardiovascular risk. Each participant received a wireless blood pressure monitor, a physical activity tracker, and instructions to download the MICArdiac app. Over a two-week period, participants were asked to measure their BP twice weekly, wear the activity tracker continuously, and engage with four weekly text messages delivered via the app. Participants completed an online daily diary for 14 days and were invited to a semi-structured interview within 7 days of the intervention period. Independent researchers with no prior relationship to participants conducted interviews, lasting 40–60 minutes, using a guide developed by the study team. Participants discussed each component of the intervention for its standalone merit and its synergistic value within the broader MICArdiac intervention. The pilot study highlighted acceptability and perceived utility, while identifying three areas requiring further refinement: (1) syncing and connectivity, (2) message personalisation, and (3) device compatibility. We addressed these concerns by: (1) developing user manuals and dedicated support staff to streamline syncing, (2) refining message content through iterative consultation with consumers, clinicians, and researchers, and supplementing messages with credible information links, and (3) expanding inclusion criteria and enhancing app compatibility with diverse wearable devices. The full details of the pilot user experience study are available in Supplement 5.

#### Control group

The control arm will receive a standard text message program, similar to published trials TEXTME^29^ and TextMe2^28^, which demonstrated efficacy in improving modifiable CVD risk factors. Messages will be randomly selected from 5 topic streams (general cardiovascular health, diet, physical activity, smoking, and medication adherence) and will be customised using two baseline variables (smoking status and vegetarian diet). Four messages will be sent per week and delivered as a text-message. Messages will be unidirectional, that is, participants will not be encouraged to respond to the messages and any responses provided (although monitored) will not be answered with a reply. Where there are return messages which are of a clinical nature, the message will be escalated to a medical doctor from the research team for review.

#### Outcomes and data collection

The primary outcome is the difference in mean daytime systolic BP measured by 24-hour ambulatory BP monitoring (ABPM) between intervention and control group at 6 months. Secondary outcomes include: 24h mean systolic and diastolic BP; first systolic and diastolic BP measured by ABPM (equating to office BP); mean daytime diastolic BP; night time mean systolic and diastolic BP (all measured by ABPM); controlled SBP (daytime SBP below 135 mm Hg daytime average ABPM); LDL cholesterol, total cholesterol, HDL cholesterol and triglycerides; controlled LDL cholesterol (under 1.8 mmol/L); BMI; diet (fruit and vegetable intake); physical activity; smoking cessation; alcohol intake; total number of risk factors controlled (systolic BP, LDL, BMI, diet, physical activity, smoking, alcohol); hypertension knowledge; GP visits; BP medication up-titration; cholesterol lowering medication up-titration.

**Table 1.** Study outcomes and collection time points.

| **Outcome** | **Measure** |
| --- | --- |
| SBP, DBP | Average SBP and DBP measured using 24h ABPM at baseline and 6 months post-randomisation (first, diurnal, nocturnal, overall). |
| Fasting LDL-C, TC, HDL-C, Triglycerides (mmol/L) | Fasting blood sample at baseline (or done in last 30 days) and 6 months post-randomisation. |
| BMI | Self-reported weight and height to calculate BMI at baseline and 6 months post-randomisation. |
| Diet (Vegetables and fruit consumption) | Self-reported: Validated Single-Question for fruit intake and for vegetable intake<sup>30</sup> at baseline and 6 months post-randomisation |
| Physical activity | Self-reported at baseline and 6 months post-randomisation |
| Smoking cessation | Self-reported smoking status at baseline and 6 months post-randomisation |
| Alcohol use | Self-reported alcohol use at baseline and 6 months post-randomisation |
| Total number of risk factors controlled | Guideline-controlled SBP, LDL, BMI, diet, physical activity, smoking, alcohol at baseline and 6 months post-randomisation |
| Hypertension knowledge | Self-reported — Modified version of Hypertension Evaluation of Lifestyle and Management Knowledge Scale<sup>31</sup> at baseline and 6 months post-randomisation |
| GP visits | Self-reported number of primary care visits, in the past 6 months, collected at baseline and 6 months post-randomisation. |
| Medication up-titration | Self-reported BP medication usage, medication class, medication dosage at baseline and 6 months post-randomisation |
| Acceptability and experience | Questionnaire administered at 6 months |

There will be two laboratory data collection timepoints: one at baseline and another post-delivery of the 6-month program. At both time points ABPM and lipid profile blood test assessments can be completed at community pathology sites using a pathology referral form provided to the participant; participants will also have the option of completing the ABPM with a study-provided machine, either fitted by a research assistant on site or mailed to the participant with instructions and available phone support. All ABPM machines used in the study are validated (community pathology laboratories use the IEM Mobile-O-Graph Self-reported and the Norav NBP-24 NG; study-owned machines are the Suntech Oscar 2 and Somnomedics ABPM Pro). Participant surveys will be completed remotely via a web link. At baseline and 6 months assessments, a research assistant will collect information including medical history and medication data from participants, in person or via the phone. Eligibility criteria, medical history and health literacy (BRIEF health literacy survey^32^) will be collected at time of registration. In addition, participants will complete questionnaires to self-report on lifestyle outcomes and healthcare service utilisation at baseline and 6 month follow up. Participants in the intervention group will be invited to participant in the process evaluation. Full details of the outcome data collected are presented in Table 1.

A **process evaluation** will be conducted following the Medical Research Council framework^33^ to assess feasibility, engagement, and implementation, through the use of *intervention usage metrics*, *online questionnaire*, and *semi-structured interviews* with participants. Surveys and interviews will explore acceptability, usability, barriers and enablers to engagement; user preferences regarding personalised messages; and patient perspectives on the use of AI for optimisation of educational content delivery.

**Intervention usage metrics** will be automatically collected via a time-stamped electronic log recording app usage (e.g., frequency and duration of use of app features and overall app use; messages received and liked; data collected from wireless monitoring devices). Additionally, a log will be kept of responses received from participants and when participants contact the study team, including the reason for contact and the method used.

To examine acceptability and feasibility of MICArdiac, all participants in the intervention group at the 6-month follow up period will be asked to complete a structured **online questionnaire** about their experiences of being sent health text messages customised using wireless device data and machine learning. Questions will explore participants’ acceptability, preferences and understanding regarding the messages (messages they remember, liked or disliked, whether messages were shown to family/friends, whether the messages prompted some sort of action or behaviour change, whether messages were well understood or confusing). Questions will also explore aspects related to intrusiveness, timing and content suitability of text messages, as well as any potential concerns regarding privacy.

Furthermore, a subset of participants in the intervention group will be invited to participate in **semi-structured interviews**. During the interviews, participants will be asked to elaborate on their previous responses to the online questionnaire, particularly exploring what they liked or disliked about the program, perceived utility and impact on their health behaviours and overall health, barriers and enablers to engagement; user preferences regarding personalised messages; and perspectives on the use of AI for lowering BP and improve cardiovascular health. These participants will be purposefully selected to diversify the opinions and views by choosing participants with ethnically, culturally, and socioeconomically diverse backgrounds, as well as with different levels of engagement with the intervention. We aim to conduct a minimum of twenty interviews, with data analysis occurring in parallel and additional individuals being recruited until there is the perception of information redundancy relative to our research objective^34–41^. Depending on participant preference, interviews will be conducted online, via telephone, or at Westmead hospital or University of Sydney affiliated meeting space and participants will be reimbursed for their travel and parking expenses. Interviews will follow a pilot tested interview guide and will be conducted by a trained interviewer.

Interviews will be digitally recorded and transcribed.

#### Sample Size

Informed by our previous TEXTME RCT^29^, we estimated that a total of **500** participants in a 1:1 ratio (250:250) will have 96% power to detect a between-group difference in mean change in ambulatory daytime average SBP of 4 mmHg (SD 11 mmHg) using a 5% level of significance and accounting for a drop-out rate of ∼20%.

#### Recruitment

Participants are identified through cardiometabolic clinics at Westmead hospital, primary care, and the community (e.g., pharmacies, sports and religious organisations, libraries, community centres), starting in Western Sydney. Recruitment expansion to New South Wales, Australian Capital Territory, and Victoria is managed through social media advertising (e.g., Facebook, Instagram) using study-developed materials targeting a diverse audience (Figure 4) and leveraging remote trial procedures. Interested participants can register their interest online by following a link or scanning a QR-code. All potentially eligible participants referred to or who contact the study team directly undergo screening to determine eligibility to the study. If the member of the study team determines that a potential participant may be suitable for the study, a copy of the patient information sheet is provided to the potential participant and any questions are answered before the interested participant signs the consent form. Ineligible participants are informed and the reasons for ineligibility are documented in the screening log.

**Figure 4:**
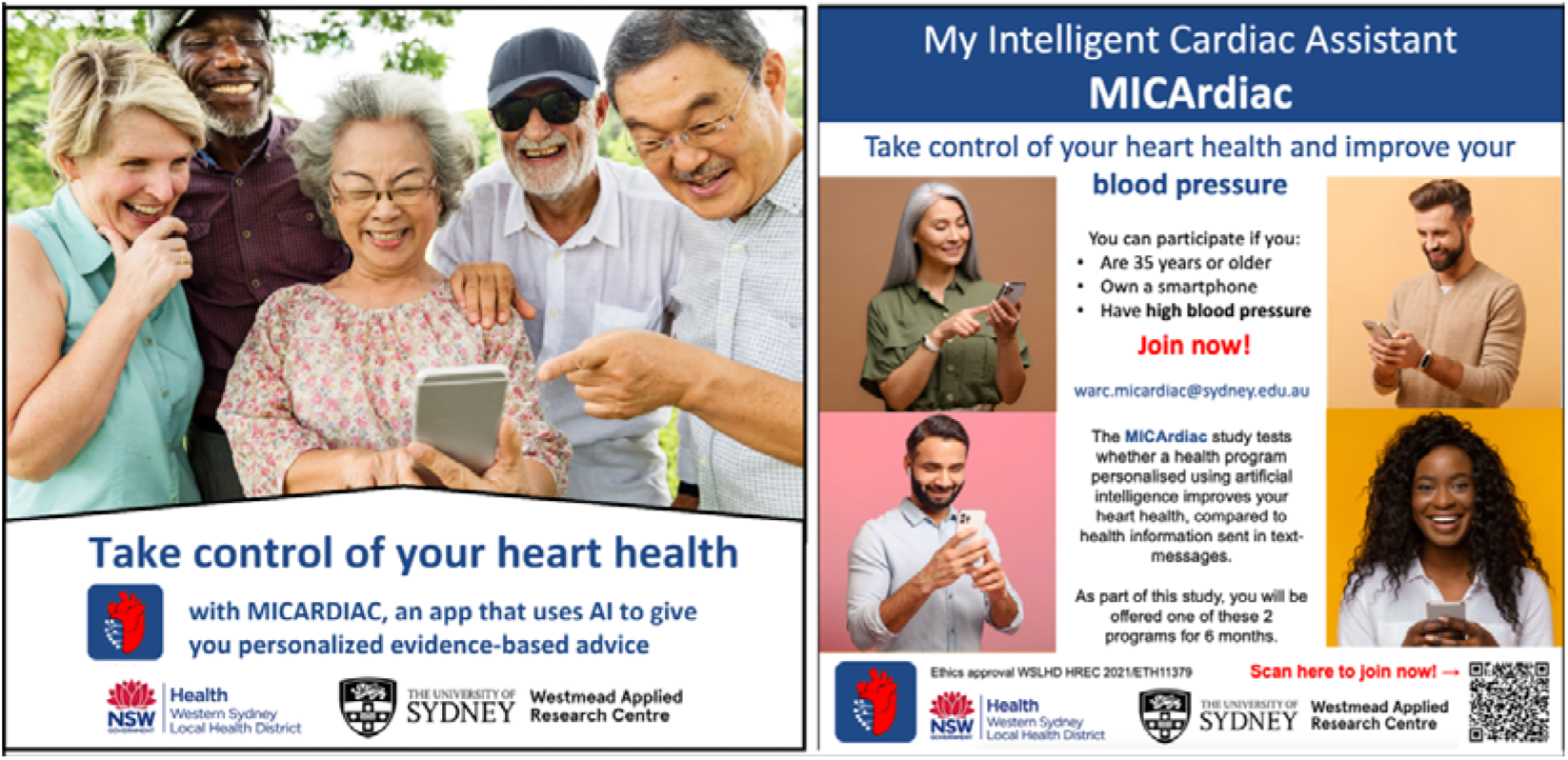
Study advertising materials for social media

#### Randomisation and blinding

Randomisation will be stratified by two variables: whether or not patients are taking blood pressure lowering medication, and by recruitment site at primary or secondary care facility. Baseline data (including objective measures and surveys) will be collected prior to randomisation. Randomisation and allocation concealment will be conducted electronically via the Research Electronic Data Capture (REDCap) software^42^. The software will automatically allocate participants to the intervention or control group, according to the randomisation sequence generated in R (using the randomiseR package) and uploaded to RedCap, ensuring allocation concealment.

Follow-up surveys will be self-filled by participants. Due to the nature of the study, participants and research study staff involved in recruitment, data collection and follow-up, will not be blinded to the randomisation outcome. However, prior to final follow-up, participants will receive a text message asking them not to reveal their allocation to study personnel until after follow-up is complete. Researchers performing data analysis will be blinded.

#### Statistical considerations and data analysis

All randomised participants will be analysed according to the intention-to-treat principle i.e. subjects will be analysed in the group (intervention or control) they are assigned to during randomisation. We will follow a published framework for intention-to-treat analysis that accounts for missing outcome data^43^: 1. Attempt to follow up all randomised participants, even if they choose to stop receiving the allocated treatment (i.e., discontinued treatment), as long as they have not withdrawn consent; 2. Perform a main analysis of all observed data that are valid under a plausible assumption about the missing data; 3. Perform sensitivity analyses to explore the effect of departures from the assumption made in the main analysis; 4. Account for all randomised participants in the sensitivity analyses.

Continuous data will be checked for normality before performing parametric tests. Appropriate non-parametric tests will be used where data are not normally distributed. All variables will be analysed by descriptive statistical methods. The number of data available and missing data, mean, median, standard deviation, interquartile range, minimum and maximum will be calculated for continuous data. Frequency tables will be generated for categorical data. For categorical variables, the number and percentage of patients with a specific level of the variable will be presented.

Outcomes at 6 months will be compared between the two groups in a regression analysis adjusting for the baseline measure of the outcome. For dichotomous outcomes we will use log-binomial regression, and for continuous outcomes we will use linear regression. Balance in baseline variables will be considered between intervention and control groups, and if significant differences exist, additional regression analysis of outcome variables adjusting for this difference will be performed.

We will report effect sizes and confidence intervals without formal adjustment for multiple comparisons, focusing on estimation rather than dichotomous hypothesis testing. All statistical analyses will be performed using R (ver. 4.3.2) (R Core Team 2020). All statistical tests will be two-tailed. P-values of less than 0.05 will be considered statistically significant unless stated otherwise. A statistical analysis plan will be finalised before data lock and unblinding.

Data from interviews will be analysed using reflexive thematic analysis^44^ of transcribed audio-recordings, using NVivo 12 (QRS International Pty Ltd., Melbourne, Australia). Themes will be identified using an inductive data-driven approach (i.e., inductive thematic analysis).^39^ The codebook will be developed and revised iteratively through discussions between researchers in the team. Identification of themes will occur by sorting the different codes into potential themes and grouping all the relevant coded data extracts within the identified themes. Themes will be identified at a semantic level, first by organizing data to show patterns in semantic content, and then by interpreting the patterns and their broader meanings and implications.^39^ After a candidate thematic ‘map’ is reached the dataset will be re-read to ensure the quality of the themes and refine them as needed. Reporting will follow the COREQ checklist for reporting qualitative research.^45^

## ETHICS AND DISSEMINATION

Ethics approval was obtained from Western Sydney Local Health District Human Ethics Research Committee (2021/ETH11379). Written and informed consent will be obtained from all study participants before commencing any study procedures. The participants will be given time to ask about the details of the study and to decide as to whether to participate in the study. Participation in the current study is entirely voluntary, and participants have the ability to withdraw at any point.

Data collected from the devices will be regularly uploaded to the iMSX Server which is linked to a cloud-based platform hosted in Australia, TextCare™, on which the MICArdiac app is based. TextCare and MICArdiac data (including data collected from wearable devices) is stored inside a Virtual Private Cloud in Amazon web services (AWS) and highly protected from unauthorised access.

### Data management

All study materials will be stored on the University of Sydney’s Research Data Store, which is a secure, enterprise-grade Network Attached Storage server located in Australia. Data for analysis (for the purposes of producing publications and presentations) will be non-identifiable data. Electronic recordings of interviews will be stored in password protected files on secure servers in University of Sydney’s Research Data Store, only accessible in original format to data manager for transcription and as source verification, and to researchers performing analyses.

### Dissemination

We aim to publish our findings in a high-reach journal to begin the process of dissemination and uptake, accompanied by a social media dissemination strategy. In addition, the findings will be presented at relevant national and international conferences.

### Patient and public involvement

The MICArdiac program was co-designed with researchers, clinicians (GP and Cardiologists) and consumers. The messages delivered by MICArdiac were co-designed and refined with consumers and healthcare professionals following published methods^46^. Patients were involved in intervention co-design, as participants to the trial and will be included in the dissemination of findings.

### Current status of the trial

Between 6 June 2023 and 1 May 2025, 451 participants have been recruited to the MICA trial. The follow-up of the last enrolled patient will be finished in November 2025. The primary results of this trial are anticipated to be available in early 2026.

## DISCUSSION

This study will evaluate the effectiveness and implementation of a novel AI-enhanced mobile health program to lower BP. It will provide a detailed assessment of uptake, engagement, and overall acceptability of this BP self-management program. The study will be limited to the Australian context; however, the cohort size is large, and recruitment will occur in the community, and primary and secondary care.

Effective BP self-management programs have generally been resourcing intensive and are not easily accessible. Our trial aims to assess a novel digital health program which is affordable, sustainable, and scalable. Successful widespread implementation of the program may have profound public health implications. Decreases in systolic blood pressure of 5 to 10 mmHg can translate into an additional 15% to 20% reduction in cardiovascular events.^47^ Our targeted lifestyle program can be conducted at relatively little expense, but if CVD can be prevented or delayed, it may result in significant reductions in morbidity and cost savings to the health system.

## Data Availability

All data produced in the present study are available upon reasonable request to the authors

## AUTHOR CONTRIBUTIONS

CKC and HK secured the funding for this project; LL, HK and CKC were involved in intervention planning and design; LL, CKC and EO developed the study protocol and statistical plan, with critical input from SM; LL, EO and CKC prepared the ethics and governance application; LL and EO prepared the first draft of this manuscript, with critical input from CKC. All authors reviewed and approved the final version of this protocol to be submitted.

## ACKNOWLEDGEMENTS

The authors would like to thank: Jim Cook and Viji Venkataramani from TechLab (The University of Sydney) and Joel Nothman, Marius Mather and the team at Sydney Informatics Hub (The University of Sydney) for their contributions in developing the AI-enhanced message optimisation algorithms. We thank Jason Chiang, Nicola Barrie, Arthur Shariev, Rahul Sathiaraj and the team at the Westmead Applied Research Centre for their support with various trial operation and administration tasks. We are grateful to Dr Peter Hay, Dr William Poh, Dr Yvette Castellino, Dr Sana Albasari, and Dr Bharat Jain for their support with participant recruitment.

## REFERENCES

1. Roth GA, Mensah GA, Johnson CO, Addolorato G, Ammirati E, Baddour LM, et al. Global burden of cardiovascular diseases and risk factors, 1990–2019: update from the GBD 2019 study. Journal of the American College of Cardiology 2020;76(25):2982–3021.

2. Murray CJ, Aravkin AY, Zheng P, Abbafati C, Abbas KM, Abbasi-Kangevari M, et al. Global burden of 87 risk factors in 204 countries and territories, 1990–2019: a systematic analysis for the Global Burden of Disease Study 2019. The Lancet 2020;396(10258):1223–1249.

3. O’Connor EA EC, Rushkin MC, et al. Behavioral Counseling Interventions to Promote a Healthy Diet and Physical Activity for Cardiovascular Disease Prevention in Adults With Cardiovascular Risk Factors: Updated Systematic Review for the U.S. Preventive Services Task Force. Evidence Synthesis, No. 195. Rockville (MD): Agency for Healthcare Research and Quality; 2020.

4. Krist AH, Davidson KW, Mangione CM, Barry MJ, Cabana M, Caughey AB, et al. Behavioral Counseling Interventions to Promote a Healthy Diet and Physical Activity for Cardiovascular Disease Prevention in Adults With Cardiovascular Risk Factors: US Preventive Services Task Force Recommendation Statement. Jama 2020;324(20):2069–2075.

5. Mancia G, Kreutz R, Brunström M, Burnier M, Grassi G, Januszewicz A, et al. 2023 ESH Guidelines for the management of arterial hypertension The Task Force for the management of arterial hypertension of the European Society of Hypertension: Endorsed by the International Society of Hypertension (ISH) and the European Renal Association (ERA). Journal of hypertension 2023;41(12):1874–2071.

6. Laranjo L, Lanas F, Sun MC, Chen DA, Hynes L, Imran TF, et al. World Heart Federation Roadmap for Secondary Prevention of Cardiovascular Disease: 2023 Update. Global heart 2024;19(1).

7. Whelton PK, Carey RM, Aronow WS, Casey DE, Collins KJ, Dennison Himmelfarb C, et al. 2017 ACC/AHA/AAPA/ABC/ACPM/AGS/APhA/ASH/ASPC/NMA/PCNA guideline for the prevention, detection, evaluation, and management of high blood pressure in adults: a report of the American College of Cardiology/American Heart Association Task Force on Clinical Practice Guidelines. Journal of the American College of Cardiology 2018;71(19):e127–e248.

8. Williams B, Mancia G, Spiering W, Agabiti Rosei E, Azizi M, Burnier M, et al. 2018 ESC/ESH Guidelines for the management of arterial hypertension: The Task Force for the management of arterial hypertension of the European Society of Cardiology (ESC) and the European Society of Hypertension (ESH). European heart journal 2018;39(33):3021-3104.

9. Virani SS, Alonso A, Aparicio HJ, Benjamin EJ, Bittencourt MS, Callaway CW, et al. Heart disease and stroke statistics—2021 update: a report from the American Heart Association. Circulation 2021;143(8):e254-e743.

10. Shahaj O, Denneny D, Schwappach A, Pearce G, Epiphaniou E, Parke HL, et al. Supporting self-management for people with hypertension: a meta-review of quantitative and qualitative systematic reviews. Journal of hypertension 2019;37(2):264–279.

11. Siopis G, Moschonis G, Eweka E, Jung J, Kwasnicka D, Asare BY-A, et al. Effectiveness, reach, uptake, and feasibility of digital health interventions for adults with hypertension: a systematic review and meta-analysis of randomised controlled trials. The Lancet Digital Health 2023;5(3):e144–e159.

12. Morawski K, Ghazinouri R, Krumme A, Lauffenburger JC, Lu Z, Durfee E, et al. Association of a smartphone application with medication adherence and blood pressure control: the MedISAFE-BP randomized clinical trial. JAMA internal medicine 2018;178(6):802–809.

13. Dorsch MP, Cornellier ML, Poggi AD, Bilgen F, Chen P, Wu C, et al. Effects of a novel contextual just-in-time mobile app intervention (LowSalt4Life) on sodium intake in adults with hypertension: pilot randomized controlled trial. JMIR mHealth and uHealth 2020;8(8):e16696.

14. Pletcher MJ, Fontil V, Modrow MF, Carton T, Chamberlain AM, Todd J, et al. Effectiveness of standard vs enhanced self-measurement of blood pressure paired with a connected smartphone application: a randomized clinical trial. JAMA internal medicine 2022;182(10):1025–1034.

15. Kario K, Nomura A, Harada N, Okura A, Nakagawa K, Tanigawa T, et al. Efficacy of a digital therapeutics system in the management of essential hypertension: the HERB-DH1 pivotal trial. European heart journal 2021;42(40):4111–4122.

16. Persell SD, Peprah YA, Lipiszko D, Lee JY, Li JJ, Ciolino JD, et al. Effect of home blood pressure monitoring via a smartphone hypertension coaching application or tracking application on adults with uncontrolled hypertension: a randomized clinical trial. JAMA network open 2020;3(3):e200255–e200255.

17. Lee P, Bubeck S, Petro J. Benefits, limits, and risks of GPT-4 as an AI chatbot for medicine. New England Journal of Medicine 2023;388(13):1233–1239.

18. Topol EJ. High-performance medicine: the convergence of human and artificial intelligence. Nature medicine 2019;25(1):44–56.

19. Muse ED, Topol EJ. Transforming the cardiometabolic disease landscape: Multimodal AI-powered approaches in prevention and management. Cell Metabolism 2024.

20. Rivera SC, Liu X, Chan A-W, Denniston AK, Calvert MJ, Ashrafian H, et al. Guidelines for clinical trial protocols for interventions involving artificial intelligence: the SPIRIT-AI extension. The Lancet Digital Health 2020;2(10):e549–e560.

21. Agarwal S, LeFevre AE, Lee J, L’engle K, Mehl G, Sinha C, et al. Guidelines for reporting of health interventions using mobile phones: mobile health (mHealth) evidence reporting and assessment (mERA) checklist. bmj 2016;352.

22. Mobile nation 2019: the 5G future: Deloitte Access Economics; 2019.

23. Michie S, Richardson M, Johnston M, Abraham C, Francis J, Hardeman W, et al. The behavior change technique taxonomy (v1) of 93 hierarchically clustered techniques: building an international consensus for the reporting of behavior change interventions. Ann Behav Med 2013;46(1):81–95.

24. Michie S, Van Stralen MM, West R. The behaviour change wheel: a new method for characterising and designing behaviour change interventions. Implementation science 2011;6(1):42.

25. Topouchian J, Zelveian P, Hakobyan Z, Gharibyan H, Asmar R. Accuracy of the withings bpm connect device for self-blood pressure measurements in general population–validation according to the association for the advancement of medical instrumentation/european society of hypertension/international organization for standardization universal standard. Vascular Health and Risk Management 2022:191–200.

26. Stergiou GS, O’Brien E, Myers M, Palatini P, Parati G. STRIDE BP: an international initiative for accurate blood pressure measurement. Journal of hypertension 2020;38(3):395–399.

27. Chow CK, Redfern J, Hillis GS, Thakkar J, Santo K, Hackett ML, et al. Effect of lifestyle-focused text messaging on risk factor modification in patients with coronary heart disease: a randomized clinical trial. JAMA 2015;314(12):1255–1263.

28. Klimis H, Thiagalingam A, McIntyre D, Marschner S, Von Huben A, Chow CK. Text messages for primary prevention of cardiovascular disease: The TextMe2 Randomised Clinical Trial. Am Heart J 2021.

29. Chow CK, Redfern J, Hillis GS, Thakkar J, Santo K, Hackett ML, et al. Effect of Lifestyle-Focused Text Messaging on Risk Factor Modification in Patients With Coronary Heart Disease: A Randomized Clinical Trial. JAMA 2015;314(12):1255–1263.

30. Cook A, Roberts K, O’Leary F, Allman-Farinelli MA. Comparison of single questions and brief questionnaire with longer validated food frequency questionnaire to assess adequate fruit and vegetable intake. Nutrition 2015;31(7-8):941–947.

31. Schapira MM, Fletcher KE, Hayes A, Eastwood D, Patterson L, Ertl K, et al. The development and validation of the hypertension evaluation of lifestyle and management knowledge scale. The Journal of Clinical Hypertension 2012;14(7):461–466.

32. Haun J, Luther S, Dodd V, Donaldson P. Measurement variation across health literacy assessments: implications for assessment selection in research and practice. J Health Commun 2012;17 Suppl 3:141–59.

33. Skivington K, Matthews L, Simpson SA, Craig P, Baird J, Blazeby JM, et al. A new framework for developing and evaluating complex interventions: update of Medical Research Council guidance. BMJ 2021;374.

34. Guest G, Bunce A, Johnson L. How many interviews are enough? An experiment with data saturation and variability. Field methods 2006;18(1):59–82.

35. Creswell JW, Poth CN. Qualitative inquiry and research design: Choosing among five approaches: Sage publications; 2016.

36. Kuzel A. Sampling in qualitative inquiry. Doing Qualitative Research. 2nd. Edited by Crabtree BF, Miller WL. In: Thousand Oaks, CA: Sage Publications; 1999.

37. Morse J. Designing funded qualitative research. InHandbook for qualitative research, ed. N. Denzin and Y. Lincoln, 220–35. Thousand Oaks, CA: Sage(1995) The significance of saturation Qualitative Health Research 1994;5:147–49.

38. Ancker JS, Benda NC, Reddy M, Unertl KM, Veinot T. Guidance for publishing qualitative research in informatics. Journal of the American Medical Informatics Association 2021.

39. Braun V, Clarke V. Using thematic analysis in psychology. Qualitative Research in Psychology 2006;3(2):77–101.

40. Saldaña J. The coding manual for qualitative researchers: sage; 2021.

41. Braun V, Clarke V. To saturate or not to saturate? Questioning data saturation as a useful concept for thematic analysis and sample-size rationales. Qualitative research in sport, exercise and health 2021;13(2):201–216.

42. Harris PA, Taylor R, Thielke R, Payne J, Gonzalez N, Conde JG. Research electronic data capture (REDCap)—A metadata-driven methodology and workflow process for providing translational research informatics support. Journal of biomedical informatics 2009;42(2):377–381.

43. White IR, Horton NJ, Carpenter J, Pocock SJ. Strategy for intention to treat analysis in randomised trials with missing outcome data. Bmj 2011;342.

44. Willig C, Stainton Rogers W. The SAGE Handbook of Qualitative Research in Psychology. London: SAGE Publications; 2017.

45. Tong A, Sainsbury P, Craig J. Consolidated criteria for reporting qualitative research (COREQ): a 32-item checklist for interviews and focus groups. Int J Qual Health Care 2007;19(6):349–357.

46. Redfern J, Thiagalingam A, Jan S, Whittaker R, Hackett M, Mooney J, et al. Development of a set of mobile phone text messages designed for prevention of recurrent cardiovascular events. European journal of preventive cardiology 2014;21(4):492–499.

47. Rahimi K, Bidel Z, Nazarzadeh M, Copland E, Canoy D, Ramakrishnan R, et al. Pharmacological blood pressure lowering for primary and secondary prevention of cardiovascular disease across different levels of blood pressure: an individual participant-level data meta-analysis. The Lancet 2021;397(10285):1625–1636.

